# Prevalence of postpartum depression and its association with Diabetes mellitus among mothers in Mbarara, southwestern Uganda

**DOI:** 10.1101/2023.10.23.23297392

**Authors:** Catherine Atuhaire, Kabanda Taseera, Esther C Atukunda, Daniel Atwine, Lynn T Matthews, Godfrey Zari Rukundo

## Abstract

**Background:** Postpartum Depression (PPD) is a major health challenge with potentially devastating maternal and physical health outcomes. Development of diabetes mellitus has been hypothesized as one the potential adverse effects of PPD among mothers in the postpartum period but this association has not been adequately studied. This study aimed at determining prevalence of postpartum depression and its association with diabetes mellitus among mothers in Mbarara District, southwestern Uganda.

**Methods:** This was a facility based cross sectional study of 309 mothers between 6^th^ week to 6^th^ month after childbirth. Using proportionate stratified consecutive sampling, mothers were enrolled from postnatal clinics of two health facilities, Mbarara Regional Referral Hospital and Bwizibwera Health center IV. PPD was diagnosed using the Mini-International Neuropsychiatric Interview (MINI 7.0.2) for the Diagnostic and Statistical Manual of Mental Disorders, 5^th^ Edition (DSM-5). Diabetes mellitus was diagnosed by measuring Hemoglobin A1c (HbA1c). Logistic regression was used to determine the association of PPD and diabetes mellitus among mothers.

**Results:** The study established that PPD prevalence among mothers of 6^th^ weeks to 6^th^ months postpartum period in Mbarara was 40.5% (95% CI: 35.1-45.1%). A statistically significant association between postpartum depression and diabetes mellitus in mothers between 6 weeks and 6 months postpartum was established. The prevalence of diabetes mellitus among mothers with PPD was 28% compared to 13.6% among mothers without PPD Mothers with PPD had 3 times higher odds of being newly diagnosed with diabetes between 6 weeks and 6 months postpartum as compared to those without PPD during the same period (aOR=3.0, 95% CI: 1.62-5.74, p=0.001).

**Conclusion and Recommendations:** Postpartum women within 6^th^ weeks to 6^th^ months have higher risks of developing diabetes mellitus. Research is needed to determine if targeted diabetes mellitus screening, prevention interventions and management will help reduce the burden.

## Background

Postpartum depression (PPD) has become a major maternal mental health challenge in both developing and developed countries^3^. In high income countries (HIC), the prevalence of PPD has been reported to be between 10-15% ^4^. However, the prevalence rate almost doubles in low-income countries (LIC). Some studies have reported a prevalence rate as high as 80% in LIC ^5^. Our recent study in southwestern Uganda revealed a PPD prevalence of 27% among six weeks’ postpartum mothers ^6^. PPD has several adverse medical and psychosocial outcomes in both the mothers and infants ^1^. PPD causes stress which makes the mother to develop negative perceptions about her child. The latter escalates the child neglect which affects the child’s psychological development thus causing intellectual deficiencies ^2^. In addition, PPD predisposes the mother to poor physical health including non-communicable diseases.

The World Health Organization (WHO) predicts that non-communicable diseases (NCDs) would account for 80 percent of the global burden of disease, causing seven out of every 10 deaths in LIC like Uganda^9^. NCDs cause about half of the premature deaths under the age of 70 ^10^. Diabetes mellitus is one of the major non – communicable diseases that causes morbidity and mortality in the world today ^7^; ^8^. Recent studies have shown that there is an increasing rate of diabetes mellitus and concurrent increase of psychological disorders like PPD among postpartum women ^11^. However, little is known about the association between PPD and the regulation of blood sugar.

Following childbirth, the sudden drop of oestrogen, progesterone and serotonin levels induces a state of distress which can manifest as PPD ^12^. Moreover, child birth itself is associated with stress and other psychosocial challenges. In order to cope, there is stimulation of the neuro-endocrine system which causes an increase in corticosteroids as well as adrenaline. These are likely to cause insulin resistance and vasoconstriction which may predispose these mothers to diabetes mellitus^12^. The depressed mother may fail to engage in desired lifestyle activities or may resort to over eating and frequent alcohol use which impairs glucose tolerance ^13^; ^14^; ^15^. In other scenarios, a woman suffering from diabetes mellitus may experience disturbance of the hypothalamic – pituitary – adrenal axis, inflammatory changes, problems in serotonergic regulation which can cause PPD ^16^; ^17^; ^18^.

Minimal studies have been carried out that relate the association of PPD to diabetes mellitus and none has been carried out in LIC like Uganda where PPD is highly prevalent. The high levels of PPD are like driven by prevailing conditions such as HIV, poverty as well as high rates of maternal and neonatal morbidity and mortality. Although it has been hypothesized that women with PPD are more likely to develop diabetes mellitus during the postpartum period, the available literature is inconclusive. There is scanty data that has looked at the development of diabetes mellitus after being diagnosed with postpartum depression. What is notable is that the number of women affected by diabetes mellitus during pregnancy and after delivery is increasing ^19^; ^20^. In 2015, it was reported that over 21 million births were affected by hyperglycaemia during pregnancy and 10-15% of these cases where within the post-partum period ^19^. This study therefore aimed at determining the prevalence of PPD and diabetes mellitus among mothers in Mbarara district and Mbarara city, southwestern Uganda.

## Methods

### Study design

It was a facility-based cross-sectional study design employing quantitative methods of data collection.

### Study setting

This study was conducted in Mbarara Regional Referral Hospital (MRRH) for the urban setting and Bwizibwera health center IV (BHCIV) in the rural setting among mothers who had come for their routine postpartum visits. The two health facilities are located in Mbarara district, southwestern Uganda. MRRH is a government owned regional referral and teaching hospital located in Mbarara City. It has a bed capacity of 350 and serves a population of over four million people in its catchment area. Every day, over 100 mothers are evaluated at the Maternal and Child Health (MCH) Unit. The MCH has sub-sections of postnatal, young child, family planning, and gynecology clinics. The Postnatal clinic has ten staff members and an average daily attendance of 30 mothers. Services offered include immunization, family planning, health education, infant growth monitoring, and examination of postpartum mothers. Approximately 650 postpartum mothers visited MRRH in the previous six months (MRRH records).

BHC IV is a health center that is based in a sub-urban region but largely serves the rural community of Kashari county, Mbarara district. It has eight midwives and an average daily attendance of 14 mothers. Approximately 350 postpartum mothers visited BHC IV in the previous six months (health facility records).

### Study population and period

Data were collected from 8^th^ February 2022 to 28^th^ July 2022. We recruited mothers from six weeks to six months’ postpartum as they visited these health facilities.

### Inclusion and exclusion criteria

Mothers were considered eligible if they had a live baby and were visiting the postnatal clinic for their 6^th^ week to 6^th^ month visit postpartum. Women were excluded if they were not the biological mother of the child brought for immunization, had a known diagnosis of diabetes mellitus in the current pregnancy, were very sick and unable to respond to questions or were taking drugs that reduce cortisol levels.

### Sample size estimation

It was determined using a formula for single population proportion with correction for a finite population ^21^. That is:

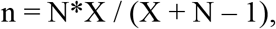

Where, X = Z_α/2_^2^ *p*(1-p) / MOE^2^,

And Z_α/2_ is the critical value of the Normal distribution at α/2 for α of 0.05= 1.96,

MOE is the margin of error considered at 5%.

p = Proportion of mothers with PPD between their 6^th^ week and 6 months post-delivery = 0.5.

N is the population size, that is, the number of postpartum mothers attending MRRH and BHC IV over a recent six months’ period = 1000.

To cater for attrition, an additional 10% was added to the sample size, hence a minimum sample size of 309 postpartum mothers was recruited.

### Sampling procedure

Using proportionate stratified consecutive sampling method, the sample size was divided proportionately among the two sites with the site as a stratum. Mothers within each site were selected consecutively based on eligibility criteria until the sample size of 309 was attained. This was based on the number of women that had given birth in each health facility the previous 6 months (hospital records).

### Study variables

The main outcome measure was diabetes mellitus among mothers clinically diagnosed with postpartum depression and the exposure variable was postpartum depression.

### Measurements

#### Hemoglobin A1c testing

To diagnose mothers with diabetes mellitus and establish their glycemic levels during the preceding 3 months, Hemoglobin A1c (HbA1c) was measured according to the standard operating procedure of the International Federation of Clinical Chemistry and Laboratory Medicine (IFCC) reference using an automated high performance liquid chromatography analyzer (Bio-Rad Diamat, Milan, Italy). The upper limit of normal was 5.6%. The reference range for HbA1c to be considered pre-diabetic was from 5.7 to 6.4% ^23^. If the HbA1C was above 6.5% then the mother was diagnosed with Diabetes mellitus based on an *International Classification of Diseases, Ninth Revision (ICD-9)* code ^24^. The results of these mothers were given to the nurse in-charges who then referred them for treatment as routine care.

### Diagnosis of postpartum depression

A mother was clinically diagnosed with postpartum depression if she was found to be positive using the Mini-International Neuropsychiatric Interview (MINI 7.0.2) for the Diagnostic and Statistical Manual of Mental Disorders, 5^th^ Edition (DSM-5) ^25^. Mothers suffering from PPD were referred for further management.

### Data collection Procedure

All eligible mothers attending any of the study sites were consented and enrolled by the study nurses. They were then clinically evaluated by an experienced registered mental health clinician for PPD using the MINI according to the DSM-5 criteria. A pre-coded and pre-tested questionnaire was administered to evaluate for socio-demographic characteristics, clinical signs and symptoms of diabetes mellitus. Anthropometric measurements such as weight in kilograms using a weighing scale machine and height in meters using a stadiometer mounted against a vertical wall were also measured to the nearest 0.1kg or 0.1m respectively. The participant then had blood drawn for laboratory investigations of HbA1C.

### Data analysis

The coded data were manually checked, cleaned and double entered into EPI Info software version 7.2 by two trained database assistants. After data entry, appropriate data validation and cleaning was done prior to exporting it to STATA software version 15·0 (Stata Corp, College Station, TX, USA) for data analysis. Descriptive analysis of participants’ characteristics was done and descriptive statistics were presented as means and standard deviations for continuous variables and frequency and percentages for categorical variables. The prevalence of PPD was calculated as a proportion of postpartum mothers diagnosed with postpartum depression using the MINI out of all mothers enrolled into the study and expressed as a percentage with its corresponding 95% confidence interval (CI).

The proportion of mothers with diabetes mellitus among those with PPD was calculated as a fraction of mothers diagnosed with diabetes mellitus out of all mothers enrolled into the study with a PPD diagnosis and was expressed as a percentage. In the same way, the proportion of mothers with diabetes mellitus among those without PPD was calculated as a fraction of mothers diagnosed with diabetes mellitus out of all mothers enrolled into the study and but had no PPD diagnosis and was also expressed as a percentage. Using Pearson Chi-square test, the two percentages were compared so as to establish if there was a significant difference in the prevalence of diabetes mellitus among mothers with and without PPD. A significance level of 5% was considered.

Bivariate analysis was done to evaluate the associations between independent (PPD) and dependent (diabetes mellitus) variables using chi-square and logistic regression. A similar analysis was performed on other maternal factors (independent variables) to establish their association with diabetes mellitus and identify possible confounders to adjust in the relationship between PPD and diabetes mellitus. Variables which were found to be statistically significant in the bivariate analysis (associated with *p* value ≤ 0.05), those with p<0.1 and those with biological plausibility (age) with regard to diabetes mellitus were included in the multivariable model building that utilizing a manual backward stepwise elimination method. PPD was locked in the model for its effect on diabetes mellitus to be adjusted for other confounders. In addition to PPD, all other variables in the final model were reported together with their adjusted OR with the corresponding 95% confidence intervals. A significance level of 5% was considered in this analysis.

## Results

We enrolled a total of 309 mothers who had a live baby and were visiting the postnatal clinic for their 6^th^ week to 6^th^ month postpartum period at Mbarara Regional Referral Hospital (n=185) and Bwizibwera Health Center IV (n=124). The study participants had a mean age of 26.7±5.7 years with majority aged below 35 years (88.3%), banyankore (78.3%), married (93.2%), with at least a secondary level of education (63.7%) and of Christian faith (92.2%). Slightly more than half were urban residents (57.9%) and had given birth at least twice (57%). Majority of mothers had never drunk alcohol (93.5%) as indicated in Table 1.

**Table 1:** Participants’ characteristics.

| Characteristics | n (%) |
| --- | --- |
| Health Facility |  |
| MRRH | 185 (59.9) |
| BHCIV | 124 (40.1) |
| Residence type |  |
| Rural | 130 (42.1) |
| Urban | 179 (57.9) |
| Mean age in years (SD) | 26.7 (5.7) |
| Age category in years |  |
| 17-19 | 20 (6.5) |
| 20-24 | 107 (34.6) |
| 25-34 | 146 (47.3) |
| 35-49 | 36 (11.7) |
| Parity |  |
| Primigravida (1) | 133 (43.0) |
| Low parity (2-4) | 152 (49.2) |
| Multipara (>5) | 24 (7.8) |
| Level of education |  |
| None/Primary | 112 (36.3) |
| Secondary | 103 (33.3) |
| Tertiary | 94 (30.4) |
| Marital Status |  |
| Unmarried | 21 (6.8) |
| Married | 288 (93.2) |
| Occupation |  |
| Unemployed | 77 (24.9) |
| Agricultural work | 65 (21.0) |
| Business | 126 (40.8) |
| Professional | 41 (13.3) |
| Ever drink alcohol |  |
| No | 289 (93.5) |
| Yes | 20 (6.5) |
| Participation in vigorous physical activity |  |
| No | 210 (68.0) |
| Yes | 99 (32.0) |

MRRH, Mbarara Regional Referral Hospital; BHCIV, Bwizibwera Health center IV

**Table 2:** Participants medical factors.

|  | **n (%)** |
| --- | --- |
| HIV sero-status |  |
| Positive | 27 (8.7) |
| Negative | 281 (90.9) |
| Not known | 1 (0.3) |
| Family history of depression |  |
| No | 281 (90.9) |
| Yes | 28 (9.1) |
| Family history of diabetes mellitus |  |
| No | 239 (77.3) |
| Yes | 70 (22.7) |
| Family history of hypertension |  |
| No | 212 (68.6) |
| Yes | 97 (31.4) |
| Hypertension |  |
| No | 292 (94.5) |
| Yes | 17 (5.5) |

### Prevalence of postpartum depression diabetes mellitus

Of the 309 mothers enrolled in the study, 125 were diagnosed with PPD based on the DSM-5, hence a prevalence of 40.5% (95% CI: 35.1-45.1%). We noted an increasing trend in prevalence of PPD with age but was not statistically significant, p=0.337. See table 3.

**Table 3:**
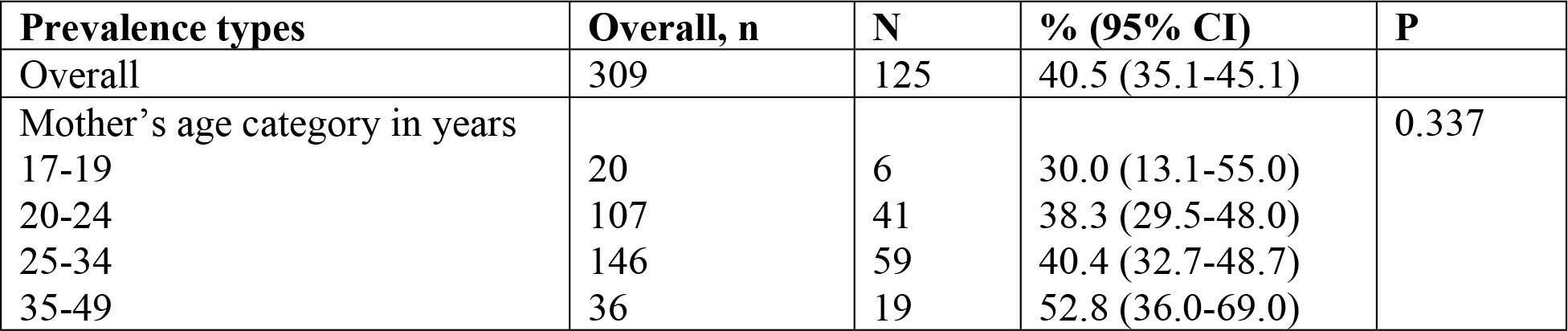
Prevalence of postpartum depression in mothers between 6 weeks and 6 months postpartum.

### Association between postpartum depression and diabetes mellitus in mothers between 6 weeks and 6 months postpartum in Mbarara district, Uganda

Of the 309 enrolled mothers, 60 were diagnosed with diabetes during postnatal visits between 6 weeks and 6 months postpartum, giving a prevalence of diabetes of 19.4% (95% CI: 15.4-24.2%). Diabetes was diagnosed in 28% (35/125) versus 13.6% (25/184) of mothers with and without PPD respectively, and this difference was statistically significant, *p=0*.*002*.

In bivariate analysis, postpartum depression was significantly associated with the diagnosis of diabetes mellitus in mothers between 6 weeks and 6 months postpartum (OR=2.5, 95% CI:1.39-4.39, p=0.0018). At multivariate analysis, PPD remained significantly associated with diabetes mellitus diagnosis after adjusting for health facility, education, age and HIV serostatus. Mothers with PPD had 3 times higher odds of being diagnosed with diabetes mellitus between 6 weeks and 6 months postpartum as compared to those without PPD during the same period after adjusting for other factors in the multivariable analysis model, (OR=3.0, 95% CI: 1.62-5.74, p=0.001) as shown in Table 4.

**Table 4:**
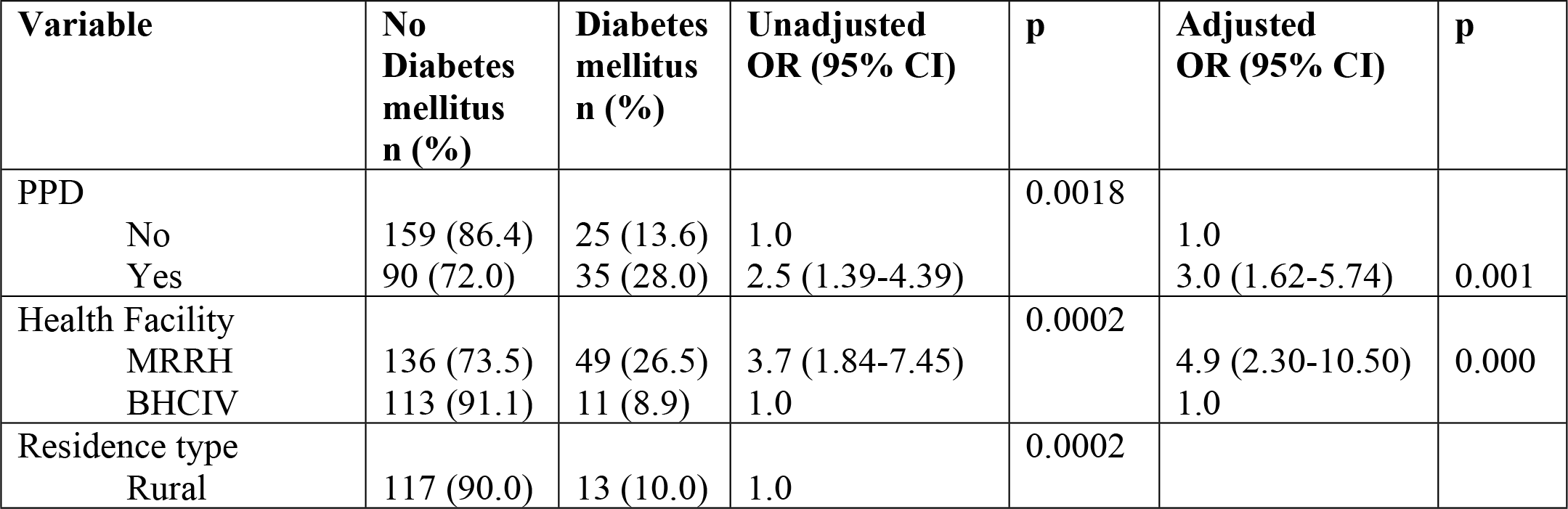
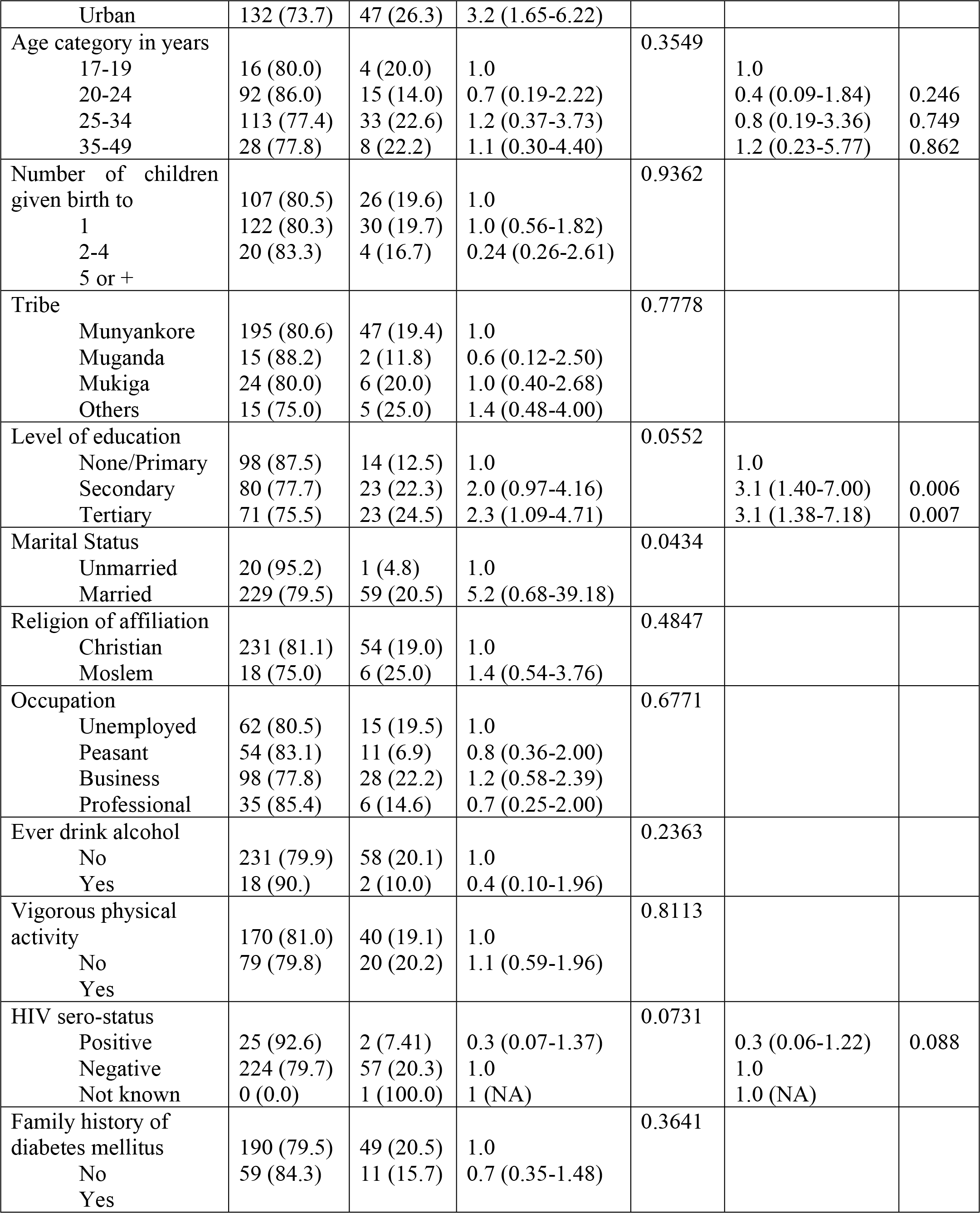
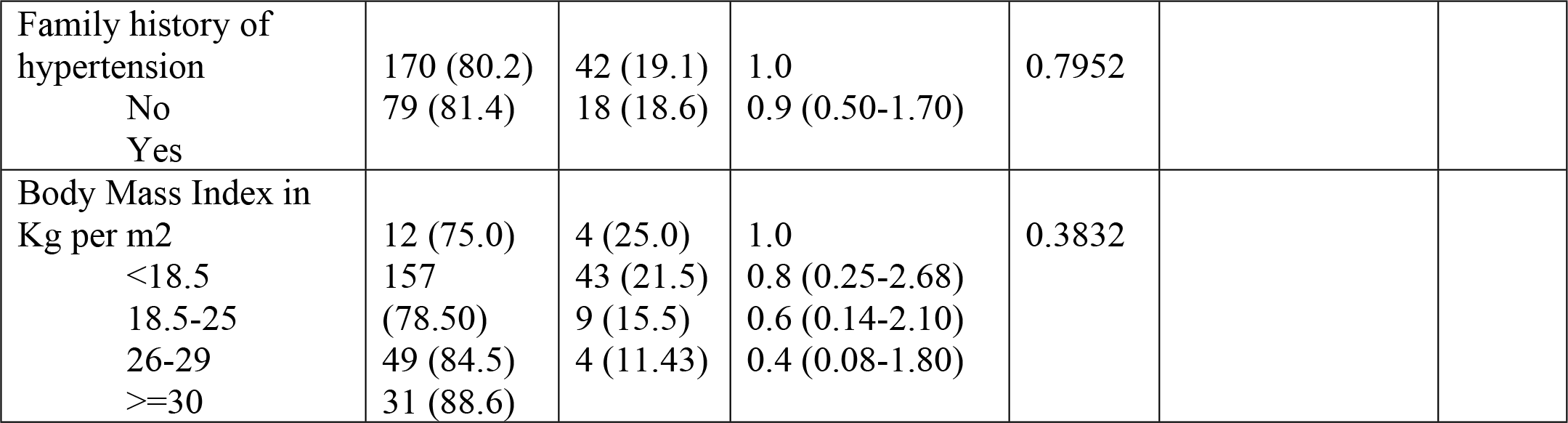
Results of bivariate and multivariate analysis for the association between postpartum depression and diabetes mellitus, and also with other factors in mothers between 6 weeks and 6 months postpartum in Mbarara district, Uganda.

Other factors that showed an independent significant association with diabetes mellitus were Health facility level where the mother was getting postnatal care (p<0.001) and having at least a secondary level of education (p<0.01). Although marital status was significantly associated with diabetes mellitus in bivariate analysis, it lost its association in multivariate analysis and was excluded from the final model.

Mothers receiving postnatal services at a regional referral hospital of Mbarara had 4.9 times higher odds of being diagnosed with diabetes mellitus as compared to those at a health center IV in Bwizibwera, aOR=4.9, (95% CI:2.30-10.50). Also, mothers having at least a secondary level of education and above had 3 times higher odds of being diagnosed with diabetes mellitus as compared to those with no or primary education (See Table 4).

## Discussion

This study aimed at determining prevalence of postpartum depression and its association with diabetes mellitus among mothers in Mbarara District, southwestern Uganda. Of the 309 mothers enrolled in the study, 125 were diagnosed with PPD based on the DSM-5, hence a prevalence of 40.5%. Similarly, the prevalence of diabetes mellitus was 19.4%. Diabetes mellitus was diagnosed in 28% of the mothers with PPD compared to 13.6% among mothers without PPD.

### Association between postpartum depression and diabetes mellitus in mothers between 6 weeks and 6 months postpartum

We observed a statistically significant association between postpartum depression and diabetes mellitus in mothers between 6 weeks and 6 months postpartum. The prevalence of diabetes mellitus among mothers with PPD was 28% compared to 14% among those without PPD. It implies that prevalence was 2.5 times higher among mothers with PPD compared to the mothers without PPD at bivariate level. After adjusting for other associated factors, the study found that mothers with PPD had 3 times higher odds of being diagnosed with diabetes mellitus between 6 weeks and 6 months postpartum as compared to those without PPD during the same period. This is so because during pregnancy and after child birth, there is a dramatic drop in the production of estrogen and progesterone hormones ^12^ which leads to postpartum depression. Studies revealed that as mothers try to cope with stress associated with childbirth, there is stimulation of the neuro endocrine system thereby increasing corticosteroids and adrenaline which leads to insulin resistance and vasoconstriction ^12^. It is also argued that during the postpartum period, mothers are likely to be withdrawn from their desired lifestyle activities or to adopt to high risky activities such as over eating that impair glucose tolerance ^13^; ^14^; ^15^.

There are generally few studies to refer to in order to collaborate and validate the findings of this study as a big proportion of the available literature have focused on gestational diabetes mellitus during pregnancy as a risk factor of depression during pregnancy and postpartum period. Literature on the association between PPD and diabetes mellitus mellitus is largely missing. However, a few studies have revealed the existence of a bidirectional relationship between postpartum depression and diabetes mellitus among mothers ^35^. Studies reveal that mothers with postpartum depression or other depressive disorders during pregnancy and postpartum period had a high risk of developing diabetes mellitus ^35^; ^36^. Similarly, in a study conducted to assess the risk factors for postpartum depression, results showed that gestational diabetes mellitus was strongly associated with increased risk for PPD in women regardless of their depression history (n=707,701) while pre-gestational (type 1 and type 2) diabetes mellitus represented an increased risk only in women with a history of depression ^32^. Also in a study conducted to examine the association between diabetes mellitus and perinatal depression among mothers from LICs (n=11,024) who were continuously enrolled in Medic aid for 6 months prior to delivery and 1 year after giving birth, results showed that after controlling for the effects of age, race, year of delivery and perinatal birth, women with diabetes mellitus had double odds of experiencing PPD during the perinatal period compared to those who were not diabetic. ^37^. All these studies comply with the study findings, because they have been conducted in similar populations who were either pregnant women or women who had given birth within one years’ time. Contrary to study findings, in a study by ^38^, results showed that in the first year postpartum, there was no association between diabetes mellitus and depression. However, after first year postpartum, women with diabetes mellitus had a nearly twofold greater risk of being diagnosed with depression compared to those without diabetes mellitus during the prenatal period.

### Prevalence of postpartum depression based on the DSM-5

The present study clinically diagnosed PPD among mothers of 6^th^ week to 6^th^ month postpartum period using the DSM-5. It reported a prevalence of 40.5% (95% CI: 35.1-45.1%). The prevalence rate reported by the present study is much higher than prevalence of 27.1% (95% CI: 22.2-32.5) previously reported in the same area of study which also used the same method of diagnosis ^6^. The difference in prevalence from the previous study is largely attributed to the different postpartum period considered by the two studies. While the current study considered a postpartum period of 6^th^ week to 6^th^ month, the previous study looked at 6 weeks postpartum. Studies from other countries in Africa have also shown discrepancies in PPD prevalence. One study carried out in Nigeria among six weeks postpartum mothers reported a prevalence of 35.6% which is in same range as the present study ^26^. The highest reported prevalence of 61.8% and 50.4% within 12 months postpartum were reported in Lime, Cameroon and DRC respectively ^27^; ^28^. Generally, studies carried out in African countries such as South Africa, Ethiopia, Zimbabwe and Cameroon have reported varying but lower rates of 31.7%, 22.9%, 33% and 31% respectively ^29^; ^30^; ^31^. Variations in timeframes used in diagnosis of PPD by numerous studies remains the main explanation for variances in prevalence rates reported. It is argued that a longer postpartum period leads to increased exposure of mothers to other risk factors of depression other than childbirth explaining the higher prevalence. Variations in the PPD prevalence is also attributed to varying tools of diagnosis and cut-off points, differences in geographical locations, study designs and units of analysis. Study results imply that mothers between 6^th^ weeks to 6^th^ months after childbirth in Mbarara district had a high risk of developing postpartum depression.

Additionally, study results also show that the risk of PPD increases with increase in age of the mothers. Study results are in support of the study carried out by ^32^ and ^33^ which also found out that the risk of PPD increased with advanced age whereby mothers aged 30 years and above were at a higher risk of PPD as compared to those aged below 30 years. Contrary to study the findings, ^34^ found out that adolescents were at a higher risk for PPD. Study results may not be similar to that done by Lanzi et al because these studies were conducted in different populations.

### Other factors associated with diabetes mellitus in mothers between 6 weeks and 6 months postpartum in Mbarara district, Uganda

This study also examined the association between diabetes mellitus and socio-demographic characteristics and medical factors. Socio-demographic characteristics examined included age, residence, marital status, education level, health facility attended, number of children, occupation of mother, physical activity and alcohol drinking habits. Medical factors examined were HIV sero-status, having a family history of hypertension, depression or diabetes mellitus, BMI and being hypertensive. The multivariate analysis revealed that two other factors were associated with being diagnosed with diabetes mellitus between 6 weeks and 6 months postpartum. The study reported having at least a secondary level of education to be independently associated with diabetes mellitus (p<0.01) agreeing with the findings of another study of 373 postpartum gestational diabetes mellitus mellitus patients carried out at a major academic centre in USA that reported an OR of 2.77 (95% CI: 09-9.00) for patients with high school education when compared to those with advanced degrees ^39^. This implies that education level affects the awareness and management of diabetes mellitus in that mothers with higher education levels are exposed to websites, reading materials and other sources that can provide all the relevant information about the preventive and management ways of diabetes mellitus mellitus hence reducing the chances of developing diabetes mellitus. However, the reverse is true for mothers with low education level. Also Health facility level was independently associated at p<0.001) whereby women in Mbarara Regional Referral Hospital (MRRH) had 4.9 increased odds of being diagnosed with diabetes mellitus as compared to women in HCIV in Bwizibwera. Health facility level was found out to be associated with diabetes mellitus mellitus because the level of a health facility in Uganda corresponds with the nature of health services being offered at that particular health facility. In this case, women who had gone in MRRH had increased odds because there are enough equipment and health care personnel to examine these women unlike when it comes to HCIVs which provide a limited number of services, are not well equipped with adequate health care staff and the necessary equipment as well.

### Strength and Limitations

The main strength of the study is the use of Mini-International Neuropsychiatric Interview (MINI 7.0.2) for the Diagnostic and Statistical Manual of Mental Disorders, 5^th^ Edition (DSM-5) and Hemoglobin A1c (HbA1c) which scientifically recognized and validated methods of diagnosing PPD and diabetes mellitus respectively. Additionally, covering a wider postpartum period of between 6 weeks and 6 months was also important in ensuring the problem is not under reported. The major weakness of the study is that the participants were selected from only two health facilities within the radius of less than 30Km of each other which may be a limitation to the generalization of the study findings. While covering a wider postpartum period was important, the study did not capture disaggregated data on prevalence of PPD and diabetes mellitus in different postpartum periods. This makes it difficult to determine the postpartum periods with high risk of PPD and diabetes mellitus to inform targeted response.

## Conclusion

The study has concluded that Postpartum Depression is strongly associated with diabetes mellitus among mothers between 6 weeks and 6 months postpartum. With a prevalence of 28%, mothers diagnosed with postpartum depression are at a higher risk of developing diabetes mellitus which has negative effects on the maternal health outcomes of both the mothers and their infants. The study highlights the need for integration of diabetes mellitus prevention and management interventions in postnatal care. Also, there is need to explore the prevalence of PPD and diabetes mellitus; and their associations in a longitudinal way with a larger sample size and controlling for other metabolic factors.

### Implications

The findings of this study are very useful in improving the maternal health outcomes and attaining the Sustainable Development Goal 3 by 2040. The study provides evidence of high prevalence of PPD and diabetes mellitus among postpartum mothers and the co-existence of the two conditions can be fatal to the health of mothers and their infants. There is therefore need for interventions that will increase public awareness, screening, prevention and management of both PPD and diabetes mellitus among postpartum mothers. Health professionals such as midwives need to be trained in the management of mothers with comorbidity of PPD and diabetes mellitus mellitus. The increased capacity of health workers to detect and manage will ensure that such conditions are detected early for better case management.

### Future research

Further studies are needed looking at varying study contexts such as rural and urban, private and public health facilities and other regions of Uganda so as to have conclusive evidence on prevalence of postpartum depression and its association with Diabetes mellitus among mothers in Uganda as a whole. Without a national view, the problem of PPD and diabetes mellitus among postpartum mothers will not get the attention it deserves as it may be treated as an isolated case. Future studies may also provide disaggregated data on the association of postpartum depression and Diabetes mellitus among mothers per categorized postpartum period to help in understanding the most risk postpartum period. Such findings would be critical in ensuring targeted interventions in the context of resource constrained countries like Uganda where targeting all postpartum women may not be financially feasible. Last but not least, future studies could also determine the efficacy of targeted diabetes mellitus screening, prevention interventions and management in reducing the burden among postpartum mothers.

## Data Availability

All relevant data are within the manuscript and its Supporting Information files

## List of abbreviations

PPD: Postpartum Depression
MINI: Mini-International Neuropsychiatric Interview
HbA1c: Hemoglobin A1c
DSM: 5-Diagnostic and Statistical Manual of Mental Disorders, 5^th^ Edition
aOR: Adjusted Odd Ratio
APA: The American Psychiatric Association
UAE: United Arab Emirates, LIC-Low Income Countries
HIC: High Income Countries
WHO: World Health Organisation
NCDs: Non-Communicable Diseases
MRRH: Mbarara Regional Referral Hospital
BHCIV: Bwizibwera health center IV
MCH: Maternal and Child Health Unit
IFCC: International Federation of Clinical Chemistry and Laboratory Medicine
PI: Principle Investigator
HIV: Human Immune Virus
CI: Confidence Interval
OR: Odds Ratio

## Declarations

We hereby declare that this is an original work and has not been published elsewhere. Also, ideas of other authors that have been used in this study have been clearly acknowledged and referenced.

## Ethical approval and consent to participate

Ethical approval to conduct the study was obtained from the Mbarara University of Science and Technology Research and Ethics Committee (REF# MUREC 1/7) and the Uganda National Council for Science and Technology (HS 2673). Written informed consent to participate in the study was obtained from all participants prior to enrolment in the study, and unique identifiers were used for participant identification. All methods were carried out in accordance with relevant guidelines and regulations.

## Consent for publication

Not applicable

## Availability of data and material

All relevant data are included in the manuscript.

## Authors’ contributions

All authors made substantial contributions to conception, design, acquisition of data, or analysis and interpretation of data; took part in drafting the article or revising it critically for important intellectual content and approved the final draft. Lynn T Mathews and Godfrey Zari Rukundo are both senior authors.

## Acknowledgements

We are grateful to all mothers who participated in this research.

## References

1. Chinawa JM, Odetunde OI, Ndu IK, et al. Postpartum depression among mothers as seen in hospitals in Enugu, South-East Nigeria: an undocumented issue. Pan Afr Med J. 2016;23(1).

2. Alnabawy AAF, Ramadan IG, Marai AAF. Correlation between Postpartum Depression, Postpartum Thyroiditis and diabetes Mellitus. Ginekol Poloznictwo. 2022;17(1):1–8.

3. Liu Y, Zhang L, Guo N, Jiang H. Postpartum depression and postpartum post-traumatic stress disorder: prevalence and associated factors. BMC Psychiatry. 2021;21(1):1–11.

4. Wubetu AD, Engidaw NA, Gizachew KD. Prevalence of postpartum depression and associated factors among postnatal care attendees in Debre Berhan, Ethiopia, 2018. BMC Pregnancy Childbirth. 2020;20(1):189. doi:10.1186/s12884-020-02873-4

5. Khalifa DS, Glavin K, Bjertness E, Lien L. Postnatal depression among Sudanese women: prevalence and validation of the Edinburgh Postnatal Depression Scale at 3 months postpartum. Int J Womens Health. Published online 2015:677–684.

6. Atuhaire C, Rukundo GZ, Nambozi G, et al. Prevalence of postpartum depression and associated factors among women in Mbarara and Rwampara districts of south-western Uganda. BMC Pregnancy Childbirth. 2021;21(1):1–12.

7. World Health Organization, Public Health Agency of Canada, Canada. Public Health Agency of Canada. Preventing Chronic Diseases: A Vital Investment. World Health Organization; 2005.

8. Damé P, Cherubini K, Goveia P, et al. Depressive symptoms in women with gestational diabetes mellitus: the LINDA-Brazil study. J Diabetes Res. 2017;2017.

9. World Health Organization. Strengthening NCD service delivery through UHC benefit package: technical meeting report, Geneva, Switzerland, 14-15 July 2020. Published online 2020.

10. Islam SMS, Purnat TD, Phuong NTA, Mwingira U, Schacht K, Fröschl G. Non-Communicable Diseases (NCDs) in developing countries: a symposium report. Glob Health. 2014;10(1):1–8.

11. Koné Pefoyo AJ, Bronskill SE, Gruneir A, et al. The increasing burden and complexity of multimorbidity. BMC Public Health. 2015;15(1):1–11.

12. Lokuge S, Frey BN, Foster JA, Soares CN, Steiner M. Depression in women: windows of vulnerability and new insights into the link between estrogen and serotonin. J Clin Psychiatry. 2011;72(11):3297.

13. Snoek FJ, Bremmer MA, Hermanns N. Constructs of depression and distress in diabetes: time for an appraisal. Lancet Diabetes Endocrinol. 2015;3(6):450–460.

14. Rustad JK, Musselman DL, Nemeroff CB. The relationship of depression and diabetes: pathophysiological and treatment implications. Psychoneuroendocrinology. 2011;36(9):1276–1286.

15. Nicklas JM, Miller LJ, Zera CA, Davis RB, Levkoff SE, Seely EW. Factors associated with depressive symptoms in the early postpartum period among women with recent gestational diabetes mellitus. Matern Child Health J. 2013;17:1665–1672.

16. Schmidt CB, Voorhorst I, Van De Gaar VH, et al. Diabetes distress is associated with adverse pregnancy outcomes in women with gestational diabetes: a prospective cohort study. BMC Pregnancy Childbirth. 2019;19:1–9.

17. Azami M, Badfar G, Soleymani A, Rahmati S. The association between gestational diabetes and postpartum depression: A systematic review and meta-analysis. Diabetes Res Clin Pract. 2019;149:147–155.

18. Pawan, Sharma, Sanjay, Kalra, Yatan, Pal Singh Balhara. Postpartum Depression and Diabetes. J Pak Med Assoc. 2022;1(1):1–5.

19. Coton SJ, Nazareth I, Petersen I. A cohort study of trends in the prevalence of pregestational diabetes in pregnancy recorded in UK general practice between 1995 and 2012. BMJ Open. 2016;6(1):e009494.

20. Veeraswamy S, Vijayam B, Gupta VK, Kapur A. Gestational diabetes: the public health relevance and approach. Diabetes Res Clin Pract. 2012;97(3):350–358.

21. Daniel WW, Cross CL. Biostatistics: A Foundation for Analysis in the Health Sciences. Wiley; 2018.

22. Goyal A, Gupta Y, Singla R, Kalra S, Tandon N. American diabetes association “standards of medical care—2020 for gestational diabetes mellitus”: a critical Appraisal. Diabetes Ther. 2020;11:1639–1644.

23. Maser RE, Mitchell BD, Vinik AI, Freeman R. The association between cardiovascular autonomic neuropathy and mortality in individuals with diabetes: a meta-analysis. Diabetes Care. 2003;26(6):1895–1901.

24. World Health Organization. International classification of diseases—Ninth revision (ICD-9). Wkly Epidemiol Rec Relevé Épidémiologique Hebd. 1988;63(45):343–344.

25. Segre LS, O’Hara MW, Arndt S, Stuart S. The prevalence of postpartum depression: the relative significance of three social status indices. Soc Psychiatry Psychiatr Epidemiol. 2007;42:316–321.

26. Adeyemo E, Oluwole E, Kanma-Okafor O, Izuka O, Odeyemi K. Prevalence and predictors of postpartum depression among postnatal women in Lagos, Nigeria. Afr Health Sci. 2020;20(4):1943–1954.

27. Ghogomu G, Halle-Ekane G, Nde P, et al. Prevalence and predictors of depression among postpartum mothers in the Limbe Health District, Cameroon: a cross-sectional study. Br J Med Med Res. 2016;12(3):1–11.

28. Adama ND, Foumane P, Olen JPK, Dohbit JS, Meka ENU, Mboudou E. Prevalence and risk factors of postpartum depression in Yaounde, Cameroon. Open J Obstet Gynecol. 2015;5(11):608.

29. Hung KJ, Tomlinson M, le Roux IM, Dewing S, Chopra M, Tsai AC. Community-based prenatal screening for postpartum depression in a South African township. Int J Gynecol Obstet. 2014;126(1):74–77.

30. Toru T, Chemir F, Anand S. Magnitude of postpartum depression and associated factors among women in Mizan Aman town, Bench Maji zone, Southwest Ethiopia. BMC Pregnancy Childbirth. 2018;18:1–7.

31. Chibanda D, Mangezi W, Tshimanga M, et al. Postnatal depression by HIV status among women in Zimbabwe. J Womens Health. 2010;19(11):2071–2077.

32. Silverman ME, Reichenberg A, Savitz DA, et al. The risk factors for postpartum depression: A population-based study. Depress Anxiety. 2017;34(2):178–187.

33. Luke S, Salihu HM, Alio AP, et al. Risk factors for major antenatal depression among low-income African American women. J Womens Health. 2009;18(11):1841–1846.

34. Lanzi RG, Bert SC, Jacobs BK, Centers for the Prevention of Child Neglect. Depression among a sample of first-time adolescent and adult mothers. J Child Adolesc Psychiatr Nurs. 2009;22(4):194–202.

35. Fischer S, Morales-Suárez-Varela M. The Bidirectional Relationship between Gestational Diabetes and Depression in Pregnant Women: A Systematic Search and Review. In: Vol 11. MDPI; 2023:404.

36. Hinkle SN, Buck Louis GM, Rawal S, Zhu Y, Albert PS, Zhang C. A longitudinal study of depression and gestational diabetes in pregnancy and the postpartum period. Diabetologia. 2016;59(12):2594–2602.

37. Kozhimannil KB, Pereira MA, Harlow BL. Association between diabetes and perinatal depression among low-income mothers. Jama. 2009;301(8):842–847.

38. Pace R, Rahme E, Da Costa D, Dasgupta K. Association between gestational diabetes mellitus and depression in parents: a retrospective cohort study. Clin Epidemiol. Published online 2018:1827–1838.

39. Mathieu C, Neumann CS, Hare RD, Babiak P. A dark side of leadership: Corporate psychopathy and its influence on employee well-being and job satisfaction. Personal Individ Differ. 2014;59:83–88.

